# An Automated and Portable Platform for Rapid Cell-Free DNA Isolation and Its Application in Microbial DNA Sequencing from Human Blood Samples

**DOI:** 10.1101/2024.12.29.24319315

**Authors:** Linda Marriott, Ana Martinez-Lopez, Antonio Liga, Kazuhiro Horiba, Amanda Warr, Jacob N. Phulusa, Radhe Shantha Kumar, Laura Carey, Yoshinori Ito, Benjamin J. Parcell, Nicholas R. Leslie, Nicholas A. Feasey, Shevin T. Jacob, Jamie Rylance, Maïwenn Kersaudy-Kerhoas

**Affiliations:** School of Engineering, Heriot-Watt University, United Kingdom; The Roslin Institute, University of Edinburgh, United Kingdom; Pathogen Genomics Center, National Institute of Infectious Diseases, Japan; Malawi-Liverpool Wellcome Clinical Research Programme, Kamuzu University of Health Sciences, Blantyre, Malawi; Queens’ college, University of Cambridge, United Kingdom; Department of Clinical Sciences, Liverpool School of Tropical Medicine, United Kingdom; Department of Pediatrics, Aichi Medical University, Japan; School of Medicine, University of Dundee, United Kingdom; School of Medicine, University of St Andrews, United Kingdom

## Abstract

**Background:** The prompt identification of pathogens in human circulation in a clinically deployable format remains an unmet clinical need. The established test for infection diagnostics remains blood culture, which typically takes 2-4 days and is positive in less than 15% of cases, with many prevalent pathogens difficult or impossible to culture. While microbial cfDNA in blood could facilitate the diagnosis of sepsis and febrile and infectious conditions, sample preparation for cell-free DNA (cfDNA) analysis in decentralised settings presents challenges due to its complexity and the low concentration and fragmented nature of cfDNA in blood plasma.

**Methods:** We developed a portable and automated platform (CNASafe) for cfDNA isolation from human plasma samples. Device performance was evaluated by comparing cfDNA yield against a reference (QIAGEN QIAamp Circulating Nucleic Acid Kit). cfDNA eluates from ten non-cultured blood samples from hospital patients were sequenced on a nanopore sequencer, and results compared to blood cultures.

**Results:** Extraction of cfDNA using the CNASafe device was completed in 40 minutes, compared to the 2-hour reference protocol. The device achieved an average relative cfDNA recovery of 100.5% over 333 unique extractions encompassing all parameter variations, demonstrating a performance equivalent to the reference kit. From the patient samples, a sufficient quantity of microbial cfDNA was extracted to either identify pathogens missed by blood cultures or confirm negative cultures.

**Conclusions:** The CNASafe platform and real-time nanopore sequencing offer a promising solution for the rapid deployment of metagenomic diagnostics, enabling pathogen identification within a few hours in decentralised clinical environments.

## INTRODUCTION

Over the past few decades, circulating nucleic acids (CNAs) in blood plasma have emerged as important biomarkers for several medical applications. CNAs have been particularly well-established in oncology diagnostics (1,2), serving as an attractive alternative to conventional tissue biopsy. Tissue biopsies are not only costly and invasive but are also limited by sampling bias due to tumour heterogeneity. In contrast, liquid biopsies are less invasive and enable repeated testing for longitudinal assessments. Liquid biopsy has also been identified as a valuable predictive and prognostic tool (3), as well as an effective way to monitor treatment (4) by detecting minimal residual disease and predicting recurrence (5). Beyond personalized oncology, the potential of cell-free DNA (cfDNA) has been demonstrated across various applications. These include non-invasive prenatal testing through the detection of fetal cfDNA in maternal blood (6), organ transplantation monitoring via circulating donor DNA (7,8), and toxicology applications through circulating microRNA (miRNA) detection (9,10). More recently, cfDNA has been used to identify infectious pathogens through circulating microbial DNA detection (11–13).

In the latter case, microbial cfDNA enables the identification of disease-causing pathogens and provides accurate, rapid, and clinically actionable diagnosis of bloodstream infections. Bloodstream infections and sepsis lead to high mortality rates, prolonged hospitalizations, and pose a significant socio-economic burden globally (14). Current diagnostic procedures rely on blood cultures, which lack specificity and sensitivity, and often have a time-to-result exceeding 72 hours in clinical practice, typically after broad-spectrum antibiotics have been administered. The incorrect use of antibiotics has been associated with an increased risk of antimicrobial resistance (AMR) and mortality (15). Additionally, parasites and viruses cannot be cultured, and fungi require specific and often challenging culture conditions, limiting the utility of blood culture in infectious disease diagnosis. Protein biomarkers, such as procalcitonin (PCT) and C-reactive protein (CRP), are useful for establishing if there is an infection present but do not inform the presence of specific pathogens. PCR, including multiplex PCR panels can be effective, but have several limitations: their multiplex power is limited and, for a specific pathogen, PCR methods only probe one or a few selected genomic sites. Microbial cfDNA released from lysed pathogens in the infected human circulation has been shown to have high sensitivity for the detection of sepsis-causing pathogens, and its use could result in improvement of turnaround times (TATs) for results and improved antimicrobial stewardship (11). This is particularly valuable in conditions where the aetiology is difficult or impossible to determine through conventional methods such as blood cultures.

Whatever the clinical application considered, the hurdles are similar for liquid biopsies to reach their promise. Samples must be prepared consistently and in a way that maximises the nucleic acid yield while maintaining quality and avoiding background genomic DNA interference. CNAs are circulating in a complex matrix, comprising white blood cells, red blood cells, proteins, and metabolites, that would inhibit their detection if not removed. The current solution for isolating circulating cell-free nucleic acids from blood is a skilled, labour- and resource-intensive process, consisting of two distinct stages: (i) isolating plasma from whole blood; and (ii) isolating circulating cell-free nucleic acids from plasma. Most of the commercial methods for isolating CNAs from plasma are based on the properties of solid substrate, usually a silica membrane within a spin column to reversibly bind CNAs in presence of a binding buffer (16). QIAamp Circulating Nucleic Acid kit (Qiagen) is a market reference. However, other commercial solid phase extraction methods include the use of magnetic beads as substrate for CNA adsorption (17), such as the MagMAX cfDNA Isolation Kit from Thermo Fisher Scientific, EZ1&2 ccfDNA Kit from Qiagen, and Maxwell® RSC ccfDNA Plasma Kit from Promega. Both filtration capture columns and magnetic silica beads rely on either a manual workflow or bulky, costly, and semi-automated platforms operated by skilled staff; these approaches require a centralised lab and are not compatible with blood processing in a decentralised manner (18).

Microfluidics has emerged as a possible toolbox to miniaturise and facilitate some challenging steps of liquid biopsy analysis. The potential advantages of microfluidic platforms for point-of-care applications have been understood for many years and include full automation, self-contained reaction volumes, integration of multi-step processes, increased speed, portability, and ease of disposal (19). In the context of CNAs, some researchers focused on improving the substrate for nucleic acids adsorption, either creating microstructures (20) trapping silica beads in gel to increase surface-area (21), generating a “fluidized bed” to enhance capture on magnetic beads (22) which was later coupled to digital droplet PCR (23), or activating chip surfaces to allow CNA binding (24). While most microfluidic applications and above examples are only suitable for small volume (microlitres) samples (25), larger volume applications have been developed as well. Several platforms, including many centrifugal microfluidics have demonstrated its value for separating large volumes of whole blood, typically approaching cfDNA recovery similar to manual or robotic magnetic bead kits, on 0.5 to under 4 mL of plasma in 15 min to 2 hours, but never combining all of optimised features in a single device, and never compared to the gold standard silica membrane columns (26–30). In addition, all the above results were demonstrated for oncological applications, and none were demonstrated for clinical metagenomic applications.

Here, to address these gaps, we have developed CNASafe, a novel automated microfluidic cfDNA extraction platform. We demonstrate its use directly in iSEP-SEQ, a simple and versatile cell-free DNA metagenomic workflow to identify pathogens in non-cultured blood samples.

## MATERIAL AND METHODS

### Overall concept

The CNASafe platform comprises a consumable cartridge and an automated instrument (Figure 1A) and was designed to deliver convenient and immediate sample preparation. It requires no manual intervention in the field, such as reagent pipetting and mixing. The CNASafe platform produces CNA eluates compatible with all major analytical platforms for example quantitative PCR, sequencing workflows or biosensors (Figure 1B). CNASafe was integrated to iSEP-SEQ, an unbiased cell-free DNA (cfDNA) metagenomic workflow able to identify any DNA-based pathogen (DNA viruses, bacteria, as well as fungi and various parasites) at species level, directly from a plasma sample. iSEP-SEQ includes a rapid sequencing assay, delivering a clinically actionable result in the form of a list of top pathogens (viruses, bacteria, as well as fungi and various parasites) at species level (Figure 1C).

**Figure 1:**
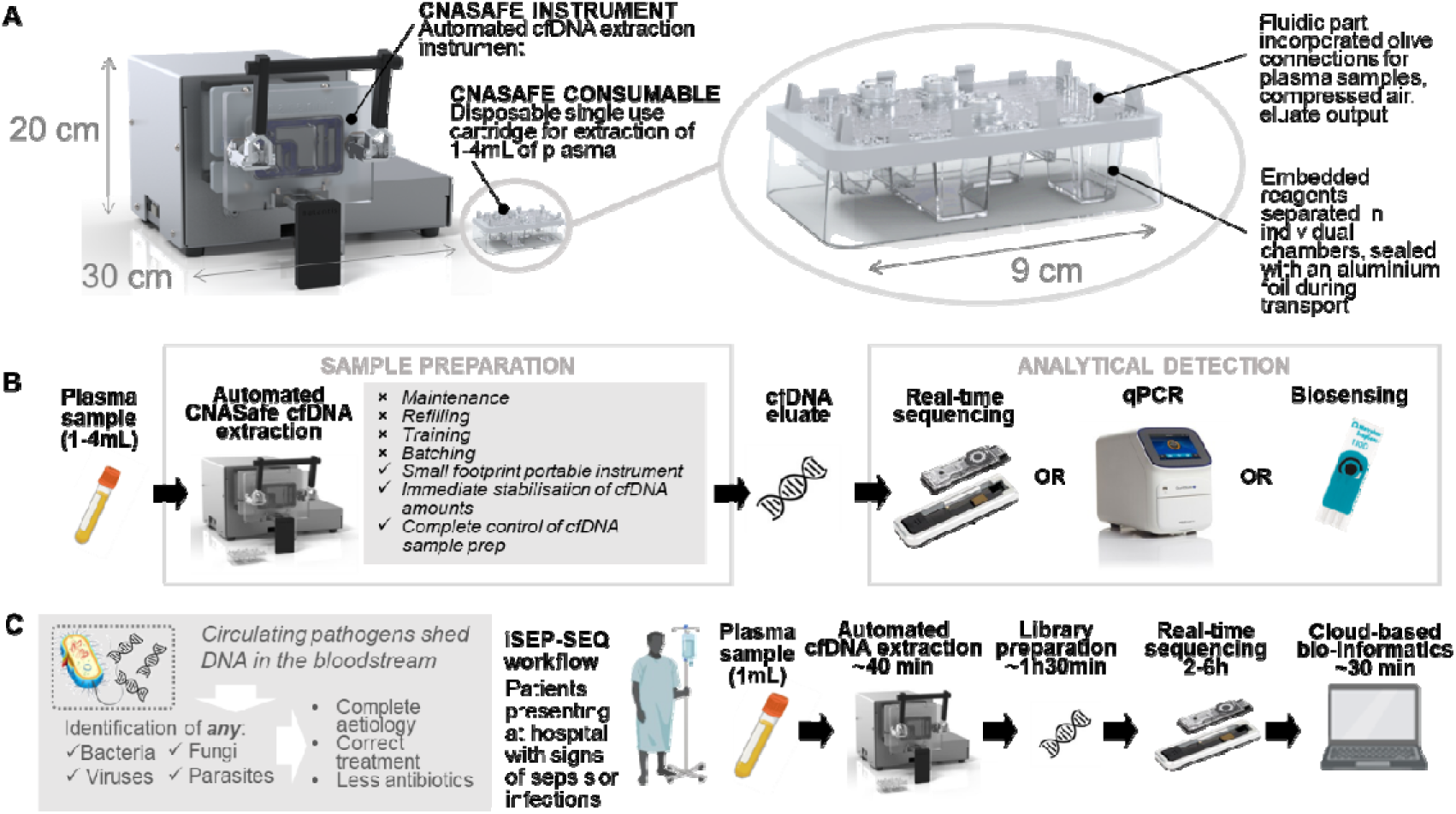
CNASafe technology overview. **(A)** CNASafe platform comprises a consumable cartridge, integrating all the reagents required for CNA extraction, and an automated instrument to successively actuate the valves integrated into the cartridge and drive the flow as needed between the reagent chambers. The platform has a small footprint suitable for decentralised and near-patient applications. **(B)** The CNASafe approach delivers cfDNA eluate compatible with numerous analytical platforms including Real-time nanopore sequencing, qPCR, and biosensors. **(C)** Overview of microbial cfDNA use and the iSEP-SEQ workflow for the identification of pathogens with CNASafe cfDNA isolation and sequencing.

### Cartridge Fabrication and Assembly

Both sides of the CNASafe cartridge were injection moulded in COC TOPAS 8007S-04 by third parties upon specified design. Further details including cartridge filling is described in Supplemental Method 1. A toggle-clamp mechanism in the instrument allows the combination of the two sides of the cartridge once placed inside the unit. Once ready to use, the sample tube is attached to the cartridge via a tube and an automated instrument. The instrument includes stepper motors to actuate the turning valves, a membrane air pump (Schwarzer precision SP270EC), and an electric heater, and is controlled by a Labview application accessible through a laptop. On completion of the workflow, the eluate obtained was stored at −20°C until use.

### Human Samples

For the CNASafe cartridge performance experiments, fresh human blood samples (Scottish National Blood Transfusion Services, contract reference 18∼20 and Ethics Approval from Office for Research Ethics Committees Northern Ireland (ORECNI) (Reference: 18/NI/0148) were spun at 3,270 g for 10 min and the plasma was removed. The plasma was then spun at 12,000 g for 10 min, removed from any pelleted material, and pooled. Aliquots of pooled plasma were stored at −20°C until further use.

For the sequencing validation with bacterial spiked samples, we created mock samples by mixing 1.5 ng of fragmented microbial community mix to 1 mL of stored human plasma (Cambridge Bioscience). The community mix containing genomes from 8 microbes (7 bacteria and 1 fungus) (ZymoBIOMICS HMW DNA Standard (D6322), Zymo Research) was fragmented by dsFragmentase (M0348, NEB) treatment according to the manufacturer’s protocol for fragments under 200 bp: 35 min incubation at 37°C followed by clean up using SPRIselect magnetic beads (Beckman Coulter).

For the iSEP-SEQ workflow validation using patient samples, frozen plasma samples from patients hospitalised with sepsis and enrolled in the Cardiovascular Responses In Septic Patients (CRISP) study were obtained through the Malawi-Liverpool-Wellcome Programme. These blood samples were obtained between October 2018 and January 2020 for blood cultures. Plasma was removed from any remaining blood samples, frozen and stored at the Queen Elizabeth Central Hospital at −80°C. Ten samples were shipped on dry-ice to Heriot-Watt University in October 2021. Upon receipt, samples were stored at −20°C until cfDNA extraction.

The CRISP study was approved by the National Health Sciences Research Committee in Malawi (Reference: 2157), and Liverpool School of Tropical Medicine Research Ethics Committee (Reference: 18-062). Approval for use in the iSEP-SEQ workflow was granted by Heriot-Watt University Research Ethics Committee (Reference: 1285 -iSEP-SEQ_ARCS / 320361).

### Blood cultures

Blood cultures were processed on an automated BacT/ALERT system (bioMérieux, France). Enterobacteriaceae and oxidase-positive Gram-negative bacilli were identified by API (BioMérieux, France). The blood culture service at the Queen Elizabeth Central Hospital is ISO15189 compliant and accredited the UK National External Quality Assessment Service (UK NEQAS). Bacteria that form part of the normal skin or oral flora, including diphtheroids, bacilli, micrococci, coagulase-negative staphylococci, and α-haemolytic streptococci (other than S pneumoniae), were considered to be contaminants.

### cfDNA manual extraction

Manual extractions were performed for benchmark references following the QIAamp Circulating Nucleic Acid Kit (Qiagen) protocol for the Purification of Circulating Nucleic Acids from Serum or Plasma (QIAamp Circulating Nucleic Acid Handbook 10/2019). cfDNA was extracted from 1-4 mL plasma, with cfDNA eluted in 65 µL Buffer AVE.

### cfDNA extractions on CNASafe device

Extractions were performed using the CNASafe cartridge. The extraction reagents used in the cartridge were adapted from the QIAamp Circulating Nucleic Acid Kit (Qiagen) in a proprietary fashion. Depending on the experimental condition (cartridge performance or workflow validation), and as individually detailed, cfDNA was extracted from 300 µL - 4 mL plasma in a total starting volume of 4 mL (volume made up to 4 mL with DPBS) and eluted in 65 −110µL Buffer AVE. The mixing time of plasma with extraction reagents (PK, Lysis and Binding buffers) varied from 3 −12 min for cartridge performance experiments and a mixing time of 3 min used for workflow validation experiments. The total extraction takes between 40 to 45 minutes for a 3 min mixing time.

### DNA quantification

For cartridge performance experiments, real-time quantitative PCR was performed using 2x Power SYBR® Green PCR Master Mix (Thermo Fisher Scientific) to amplify a 90 bp target with LINE (Long Interspersed Nuclear Element) primers (final concentration 200 nM): forward 5’-TGC CGC AAT AAA CAT ACG TG −3’ and reverse 5’-GAC CCA GCC ATC CCA TTA C-3’. 1μL of DNA template was added to a total reaction volume of 12.5 μL. A standard curve was created using Human Genomic DNA (Bioline). Thermal cycling conditions involved a 10-minute cycle at 95°C followed by 40 cycles with 15 seconds at 95°C and 60 seconds at 60°C. Samples were amplified in triplicates using Applied Biosystems StepOnePlus™ Real-Time PCR System (Applied Biosystems). A melting curve was performed in all experiments as a control measure for non-specific amplification. Quality controls are discussed in Supplemental Method 2. Absolute amounts of cfDNA in each sample (cartridge and benchmark eluates) were obtained from the standard curve. For sequencing experiments, eluates were quantified on a Qubit fluorometer (Qubit 3.0, Invitrogen, USA) using the Qubit HS assay according to manufacturer’s instructions.

### Nanopore Sequencing and Bioinformatics

For all workflow validation samples, library preparation was carried out over 120 min using the Ligation Sequencing kit (SQK-LSK110, ONT) with no modification to the manufacturer’s protocol, beyond the use of SPRIselect beads rather than AmpureXP beads. For sequencing optimisation, libraries were prepared from 50 µL of extraction eluate regardless of the DNA concentration measured using the Qubit. For sequencing of clinical samples, sequencing libraries were prepared from 45 µL of extraction eluate regardless of the DNA concentration measured using the Qubit. Libraries were sequenced on a MinION Mk1C, Minknow version 21.02.2 or greater, using single-use Flongle Flow Cells (FLO-FLG001, ONT). Sequencing was performed for 6 h with flow cell pore counts ranging from 51-95 at the start of sequencing. High accuracy basecalling was performed using Guppy version 4.0.11 or greater. Pathogen identification was carried out using the WIMP (v2021.02.10) workflow in EPI2ME (ONT) or a bioinformatic pipeline PATHDET v6.0 or greater (31). All sequencing reads generated were analysed in both the WIMP and PATHDET workflows; no trimming of adapter sequences took place before analysis. Absolute total number of reads were recorded, as well as total number of pathogens reads and number of reads for any pathogens with more than three reads. Pathogen reads were normalised as reads per million reads of total sequencing output (RPM).

### Statistical Analysis

Conditions were analysed using GraphPad Prism software. Where possible, Student’s t-test or one-way ANOVA analysis were performed. Statistical significance was determined by unpaired parametric Student’s t-test or ANOVA. Significance threshold was set at a p-value less than or equal to 0.05.

## RESULTS

### Cartridge and Platform Design and Operation

The CNASafe cartridge includes all necessary reagents and relevant fluidics to allow a fully self-contained extraction (Figure 2.A-C): one part containing all the fluidic components (Part 1), a second part storing all reagents for the solid phase extraction (Part 2), and a third part (Part 3) which acts as a transport guard and an alignment feature. There are several unique features to this cartridge arrangement: (i) the three-dimensional nature of the cartridge creates enough space for reagent storage for extraction of millilitre volumes compatible with typical clinical sample volumes for CNA applications; (ii) the reagents are embedded into double-use ‘blisters’ (i.e., needles are used to pierce and access their contents as in a traditional blister, but the rigid nature of the blisters mean that they can be used for further steps, such as mixing); and (iii) the configuration of the inlet and outlet coming into and leaving the mixing chamber enables the effective mixing of reagents followed by siphoning onto the silica membrane. The cartridge design contains an embedded silica membrane. In this work, we prioritised a cartridge connected to an external silica membrane.

**Figure 2:**
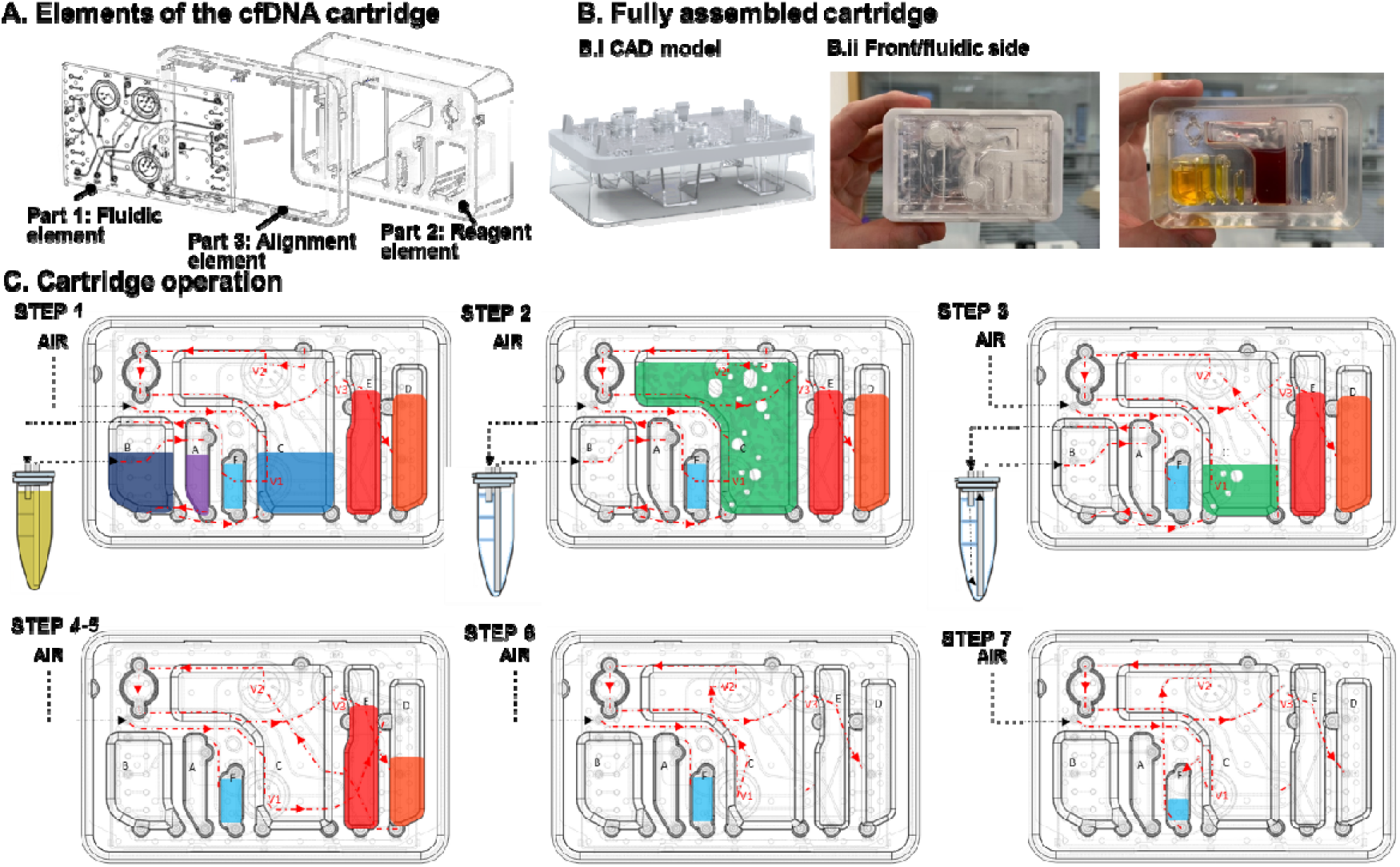
Presentation of the CNASafe consumable. **(A)** CNASafe consumable is a three-part assembly cartridge, containing a fluidic side and a reagent side, assembled with each other by an alignment frame. **(B)** B.i CAD model of the assembled cartridge B.ii. Photographs of the CNASafe cartridge, with the different reagent chambers in the back, enclosed and sealed with a foil on the front face. **(C)** Operation of the CNASafe consumable. All reagents are contained in the cartridge chambers, closed with pierceable foil. A 7-step protocol enables the cfDNA extraction from the sample tube, its retention by the silica membrane, and its elution into a collection tube for downstream characterisation. The sample is pushed by the instrument into the cartridge, where the CNA extraction occurs. Valves are indicated as V1-V3. The Chambers A-F contains respectively: Proteinase K solution, Lysis buffer, Binding buffer, Wash buffer 1, Wash buffer 2, and Elution buffer. CNAs are collected off cartridge and can be utilised for multiple assays downstream. The instrument actuates the valves, gives supply of compressed air, and is a source of heat for the drying step. In STEP 1, when the air pump is actuated, the air flow is directed towards the plasma tube, generating an overpressure which pushes the plasma within the cartridge. While doing so, Proteinase K (A) and Lysis buffer (B) are also pushed towards the Binding buffer chamber (C), which is oversized to contain simultaneously all reagents and plasma sample. In STEP 2, valves are positioned to allow the air to escape from the top of this chamber (C), while the reagents are retained at the bottom. Therefore, by actuating the membrane pump, it is possible to generate a bubble flow within the chamber, thus a turbulent flow that mixes sample and reagents. In STEP 3, valves are turned, closing the air vent and opening the liquid outlet at the bottom of the mixing chamber (C). As the air cannot escape any longer, actuating the pump generates a pressure rise within the chamber, pushing the reagent mix through the silica membrane, allowing the adsorption of nucleic acids. Liquid is collected in a waste tube. In STEPs 4 and 5, the silica membrane containing the nucleic acids is washed through a double buffer wash (D, then E). While some commercial kits use a triple wash (e.g., QIAamp Circulating Nucleic Acid Kit (Qiagen)), we opted for only two wash steps to save reagents volume, thus limiting the overall volume and weight of our disposable cartridge. In STEP 6, the silica membrane is dried for 10 min using an air flow and placing the column in a custom heating block. The automated unit includes a heater placed over the area dedicated to the membrane compartment, so this part of the protocol is fully automatable as well. Lastly, in STEP 7, the valves are turned in their last configuration and a quick burst of the air pump pushes the Elution buffer (F) to the membrane. After a 90 sec incubation, the pump is activated one last time and the eluate transferred to a collection tube for downstream use.

An automation platform to operate the cartridge was designed to take the cartridge and clamp it tight enough to bring together the three parts of the cartridge, enabling connections and thus, flow between the various compartments of the cartridge. In addition, the automation platform contains electronics to control valves and heaters enabling the control of the flow of sample and reagents within the cartridge.

Before starting the extraction protocol, the two sides of the cartridges are pressed together by the clamping system in the automation platform and a sample tube, containing 1 to 4 mL of human plasma, is connected to the cartridge fluidic side. The protocol is initiated from a laptop. FIG 2.C describes all the steps of the cfDNA extraction protocol.

### Overall extraction statistics

During the development and utilisation of the CNASafe cfDNA extraction cartridge, a total of 474 extractions were performed. The complete dataset is provided with this manuscript (https://doi.org/10.6084/m9.figshare.28075193.v1). Of these, 358 extractions were conducted after transitioning the sealing process in-house, yielding 333 successful eluates. Every single cartridge eluate was benchmarked with a manual extraction using the QIAGEN QIAamp Circulating Nucleic Acid Kit (cf Materials and methods) on the same biological sample. In all next sections, the cfDNA recovery results are reported normalised to the manual extraction reference. The average cfDNA recovery from these 333 eluates, encompassing all parameter variations, was 100.5%, demonstrating a performance equivalent to the industry gold standard for cfDNA yield. The next sections explore specific optimisation considerations.

### Influence of elution buffer volume

The first step in the optimisation of the automated workflow was to identify the elution buffer volume to be added within the CNASafe cartridge. The optimal condition should provide the maximum eluate volume, the highest cfDNA recovery, while also minimizing the volume of elution buffer (and weight) to store in the cartridge. To assess this, cfDNA was extracted from 1 mL human pooled plasma samples (mixing time 3 min), in an initial volume of 4 mL (1mL plasma + 3mL DPBS). Reagent chambers contained reagent volumes appropriate to a 4 mL starting sample. Figure 3.A represents the cartridge eluate volume extracted from the cartridge and the total cfDNA normalised to manual extraction, with the volume of elution buffer added to the cartridge: 65, 100 and 110 µL. Both the mean cartridge eluate volume and total cfDNA obtained were significantly increased with a higher elution buffer volume: a mean of 15 µL (N=15, SD=6), 43 µL (N=29, SD=7), and 53 µL (N=9, SD=10) for 65, 100, and 110 µL elution buffer volumes respectively, corresponding to an average normalised cfDNA recovery of 21, 60, and 80%. The maximum eluate volume achieved was 63 µL for 110 µL of elution buffer, for a cfDNA recovery of 67%. We did not push further the volume of elution buffer due to limitation in the maximum volume allowed in the consumable and selected 110 µL for the following experiments.

**Figure 3:**
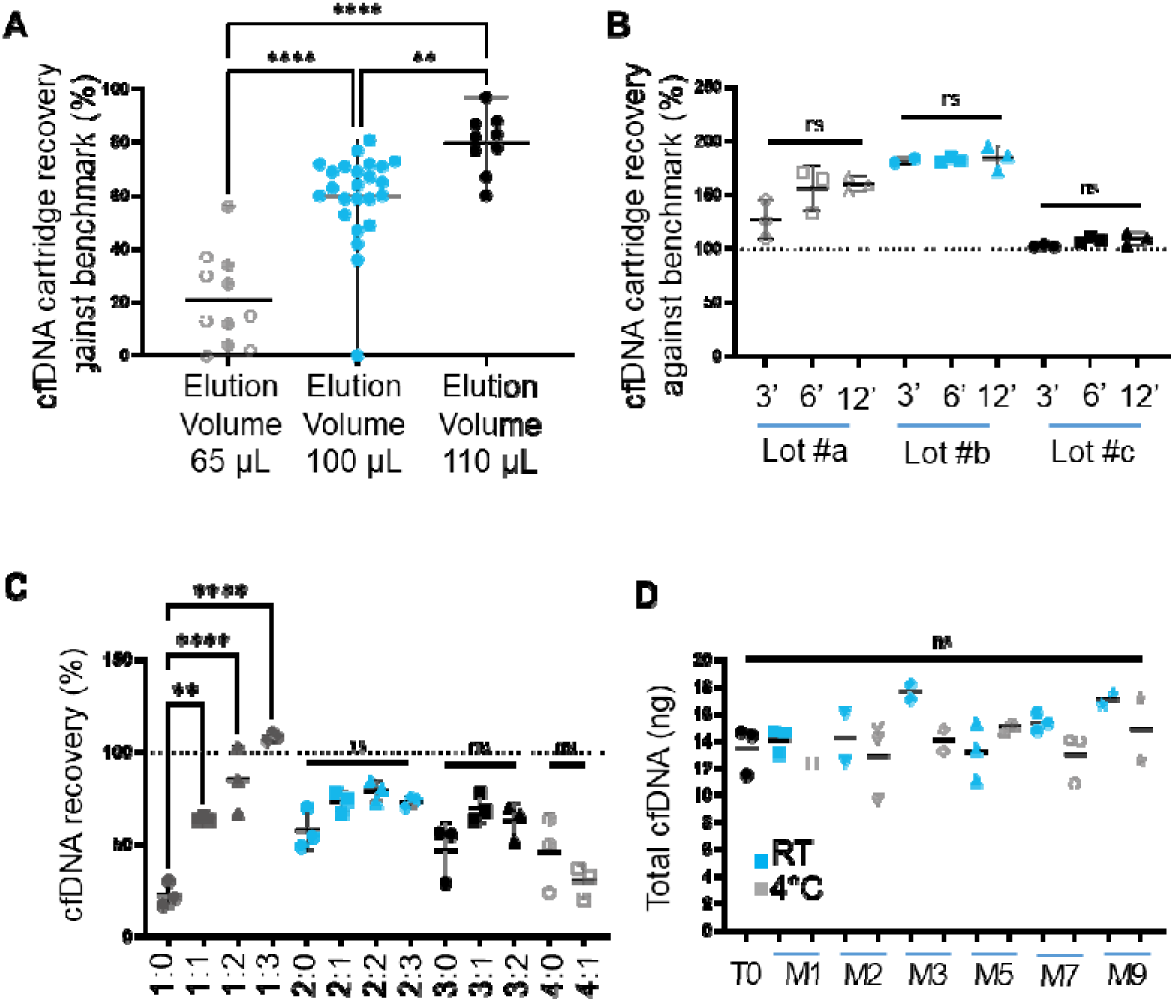
**(A)** Impact of elution buffer volume (3 min mixing) on the elution volume. **(B)** Impact of the silica membrane column and mixing time on cfDNA recovery with regards to cfDNA obtained with the manual method on the same column lot number. Lots #a, #b and #c correspond to Lots #163035846, 166014300, 172015077 according to the supplier referencing. **(C)** Impact of DPBS and plasma volume on the total amount of cfDNA recovery. **(D)** Total amount of cfDNA obtained from cartridges at different time points in months (M) following their filling and sealing, and storage conditions.

### Influence of mixing time and silica membrane lot number

The optimum sample mixing time was investigated, considering cfDNA recovery from 1 mL plasma sample in a total starting volume of 4mL (1mL Plasma + 3mL DPBS). Reagent chambers contained reagent volumes appropriate to a 4 mL starting sample. Secondly, for each sample, we compared the cfDNA extractions using the CNASafe cartridge and its silica membrane, with the manual extraction method using a membrane from the same lot and the same biological samples. Results are represented in Figure 3.B.

The QIAamp Mini columns used here were from 3 different lot numbers (#a, #b and #c). When comparing the total amount of cfDNA obtained from the extractions (not taking into account the mixing time), we observed that the column lot number had a significant impact on the result, with significant difference in the total amount of cfDNA recovered between the 3 lots (one-way ANOVA for all three column lots, for 3, 6 and 12 min, where respectively p=0.0018; p=0.0008; p<0.0001). Although the reason behind these differences has yet to be elucidated, these results highlight the importance of the silica membrane in cfDNA recovery and call for more research into the differences observed between lot numbers, and possibility to tune the design of silica membranes to improve cfDNA recovery.

The total cfDNA obtained from each mixing time was compared for each column batch: there was no significant difference seen between mixing times for lots #b and #c. Highest values were observed after 6 minutes for lot #a (9.17ng for 3min and 11.27ng for 6 min, non-significant), therefore we selected 6 min as the mixing time for the following experiments unless otherwise stated.

### Influence of plasma and DPBS sample volume

We considered 1, 2, 3 and 4 mL of plasma (mixing time 6 min) with 1-4 mL of DPBS. For all cartridge extractions performed, reagent chambers contained reagent volumes appropriate to a 4 mL starting sample. The total cfDNA amount obtained from each cartridge extraction was normalised against the manual extraction result for the same plasma volume, as illustrated in Figure 3.C.

For each volume of plasma, the highest cfDNA recovery was achieved when the total starting sample volume was equal to 4 mL. No additional increase in the recovery was observed when sample volume exceeded 4 mL, indicating that increasing the sample volume to 5 mL may have a diluting effect on the reagents. The variation of viscosity of the starting samples and its effects explain the increase of cfDNA recovery for 1 mL plasma with increasing volume of DPBS (27%, 64%, 85% and 108%, respectively for no DPBS added, 1 mL, 2 mL and 3 mL) and why the combination 1 mL plasma + 3 mL DPBS was the optimal experimental condition in terms of cfDNA recovery.

### Stability over time and temperature storage

Prefilled cartridges need to withstand a certain shelf-life in order to be stocked and used by customers when required. To determine whether the CNASafe cartridge and its reagents could withstand long-term storage, cartridges were sealed in-house and stored with the sealing foil face up, at room temperature or 4°C, for 9 months. At months 1-9, cartridges from each storage temperature were used to extract cfDNA from 1 mL of plasma in a total starting volume of 4mL (1mL Plasma + 3mL DPBS). Reagent chambers contained reagent volumes appropriate to a 4 mL starting sample. The quantity of cfDNA extracted for each condition is represented in Figure 3.D. There was no significant difference in the total amount of cfDNA obtained from the cartridges, stored at RT or 4°C, over the 9 months storage period. This confirms that the extraction reagents are stable for up to 9 months when stored in the CNASafe cartridge at room temperature or 4°C.

### Compatibility of CNASafe technology to nanopore sequencing workflow

To verify the compatibility of CNASafe cfDNA extraction with downstream nanopore sequencing, we spiked 1 mL plasma samples with 1.5 ng of fragmented microbial DNA mix (community mix of eight microbes) and extracted the cfDNA. As a comparison, we processed the same samples both with the manual reference method and with the automated CNASafe protocol using a 3 min mixing time. The eluates were then sequenced as described in the method section. Three performance indicators (classified reads, percentage of unclassified reads and Phred score) were quantified to compare benchmark (B) and cartridge (C) extractions (Supplemental Fig S3.A-C). All histograms showed no significant differences between the two extraction protocols. For all samples analysed, the percentage abundance for the eight microbes of the community mix show similar patterns between the bench and the cartridge extractions, while controls (plasma without spiked microbial mix) showed no hit (Supplemental Figure S3.D). These results confirm the compatibility of the CNASafe extraction with downstream sequencing but also indicate the capability of our integrated assay for further deployment of sepsis detection in the field. In addition, the negative controls show that the on-chip cfDNA extraction does not introduce contaminants.

### iSEP-SEQ workflow validation with human clinical samples

Following the optimisation of the sequencing workflow, the application of the iSEP-SEQ workflow to the identification of pathogens was investigated using banked plasma samples from ten patients with sepsis as described in methods. The clinical presentation of the patients is summarised in Table 1.

**Table 1:** Patient clinical data characteristics.

|  | Age | Sex | HIV/ART status | CD4 cell count (cells/uL) | Antibiotics, in the last 4 weeks | Fever | Cough | Chest pain | Headache | Vomiting | T (°C) |
| --- | --- | --- | --- | --- | --- | --- | --- | --- | --- | --- | --- |
| **CRISP 1** | 30-40 | Male | HIV+, ART | 69 | Amoxycillin | Yes | Yes | No | No | Yes | 39.2 |
| **CRISP 2** | 20-30 | Female | HIV+, ART | 240 | Erythromycin<br>Metronidazole | Yes | No | Yes | Yes | No | 38.1 |
| **CRISP 3** | 30-40 | Male | HIV+, ART | 50 | Positive, but unknown prescription | No | No | No | No | No | 38.5 |
| **CRISP 4** | 30-40 | Female | HIV+, ART | 611 | Positive, but unknown prescription | No | Yes | Yes | No | No | 36 |
| **CRISP 5** | 30-40 | Female | HIV+, ART | 45 | Ceftriaxone<br>Ciprofloxacin | Yes | Yes | Yes | No | No | 37.6 |
| **CRISP 6** | 40-50 | Female | HIV+, ART | 15 | Ciprofloxacin | Yes | Yes | No | No | Yes | 36.1 |
| **CRISP 7** | 30-40 | Male | HIV+, ART | 126 | Amoxycillin | Yes | Yes | Yes | Yes | No | 39.1 |
| **CRISP 8** | 20-30 | Male | Negative | NA | Doxycycline | Yes | Yes | Yes | No | No | 38.1 |
| **CRISP 9** | 20-30 | Female | HIV+, ART | 440 | Ceftriaxone | Yes | Yes | No | No | Yes | 34.3 |
| **CRISP 10** | 40-50 | Male | HIV+, ART | 166 | Metronidazole,<br>Bactrim | Yes | Yes | Yes | No | Yes | NA |

The samples were passed through the full iSEP-SEQ workflow. cfDNA was extracted from banked plasma samples ranging from 300 – 1000 µL using the CNASafe cartridges. All samples produced an eluate. One CNASafe extraction did not run to completion (sample 6), but an eluate from this sample was still obtained after the column was removed from the cartridge and the remaining steps (wash, dry and elution) performed on the bench. The elution volume range was 52.4±2.8 μL, excluding the sample manually recovered from the cartridge. All samples were successfully sequenced, but one set of sample files (sample 3) were lost during transfer due to an IT error. The samples were passed through *PATHDET*, a proprietary bioinformatic pipeline hosted by the University of Tokyo. In addition, *EPI2ME*, the bioinformatic platform from Oxford Nanopore Technologies was also used to compare with *PATHDET*. Statistical data from the processed samples are presented in Table 2. EPI2ME statistics are provided in Supplemental Table 1. The total number of reads varied between 47,078 and 873,892 and the percentage unclassified ranged between 5 to 12.6% (*EPI2ME* data).

**Table 2:**
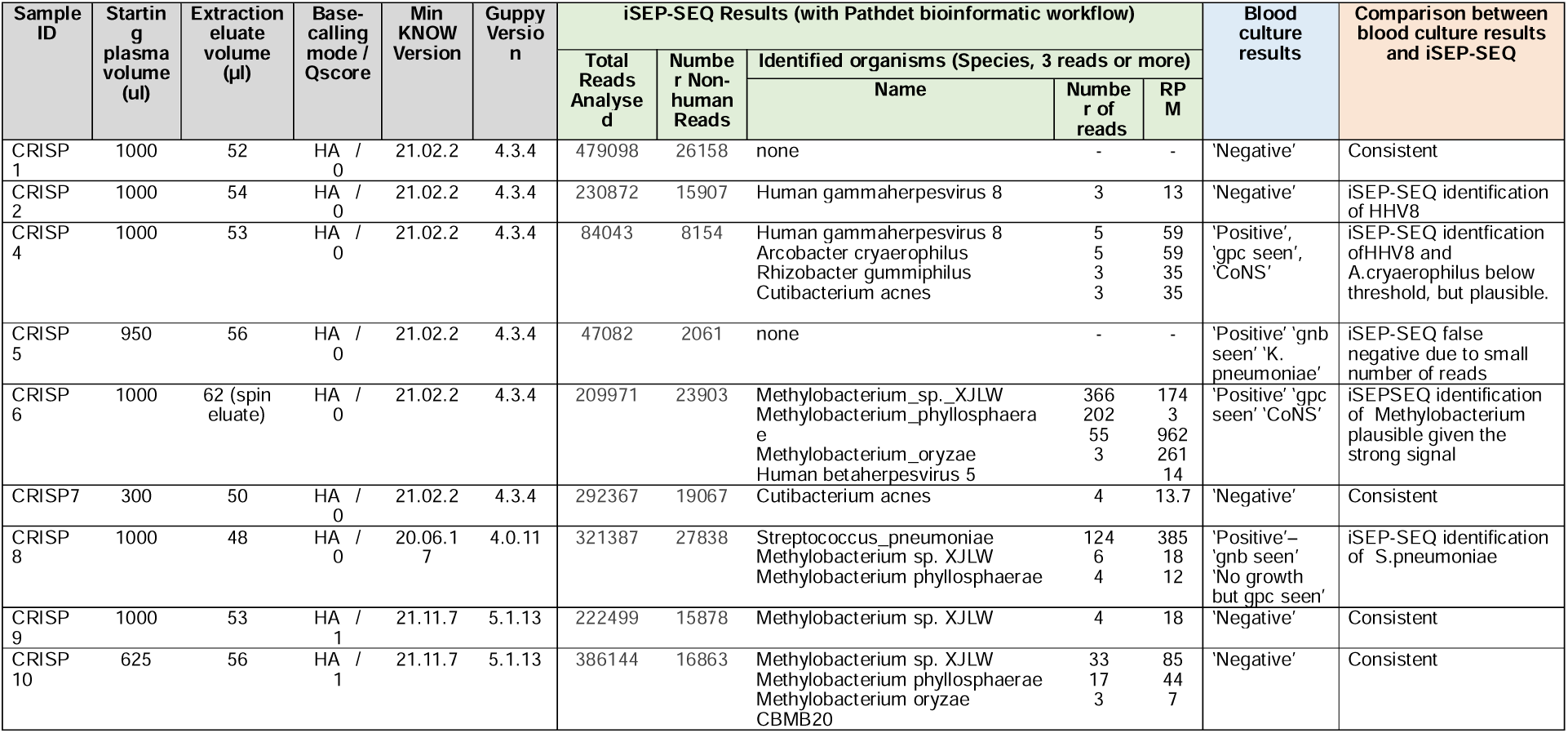
Sequencing statistics and clinical data. The blood culture results are reported as entered in CRISP study notes. CoNS = Coagulase-negative staphylococci.

In six out of nine cases, iSEP-SEQ was consistent with blood culture results. These include CRISP 1, 2, 7, 9 and 10, where the blood cultures were found negative, and where iSEP-SEQ identified bacteria in low numbers, below the threshold of 100 RPM or unculturable organisms.

In CRISP 4, iSEP-SEQ identified *Arcobacter cryaerophilus* at 59 RPM. *Arcobacter cryaerophilus* has been previously described as a pathogen in certain patients (32,33), but the signal is below the 100 RPM for positive identification of bacteria, as defined by Chiu et al (34). Human gammaherpesvirus 8 (HHV-8) was also found at a level of 59 RPM, above the threshold generally set for viruses (35). This virus would not have been identified in the blood culture. Although not a sepsis-causing pathogen, HHV viruses are part of the cytomegaloviruses (CMVs) and can cause community-acquired pneumonia (CAP) in compromised hosts (36) and a major cause of morbidity in patients with AIDS and other immunosuppressive disorders (37). The patient’s severe dyspnoea, chest pain and cough with mucus support this possibility.

In CRISP 5, cultures identified *K. pneumoniae*. *K.pneumoniae* is consistent with the presentation of the patient, and likely to have been missed by the iSEP-SEQ assay due to a lower number of reads in that particular sample. This could be prevented in the future by processing larger volumes of plasma.

In CRISP 6, iSEP-SEQ identified three Methylobacterium species which cultures failed to detect, and instead, blood cultures identified coagulase-negative staphylococci, which is considered to be a contaminant from the patient’s skin. Methylobacterium is hard to grow in culture and could have been present in the samples, but not visible on cultures.

Methylobacterium is a Gram-negative, aerobic bacterium which exists in soil, dust, water, and sewage, and rarely cause human disease, but have been previously identified as responsible for bacteremia in immunocompromised patients, such as patients with HIV included in this analysis (38,39). It is also a known contaminant of DNA extraction kits (40); however, the high RPM value (2,966 RPM at Methylobacterium genus level) and the fact that another sample was extracted after CRISP 6 using reagents from the same potentially contaminated batch number and showed no evidence of such heavy Methylobacterium contamination, point to this being a true positive.

In CRISP 8, iSEP-SEQ workflow identified a strong *Streptococcus pneumoniae* signal (124 reads corresponding to 8% of total reads and covering 9% of the organism’s genome). This was not identified in blood culture, which only stated that gram-positive cocci was observed. *Streptococcus pneumoniae* is a bacteria responsible for severe lung infection, a result highly consistent with the presentation of the patient (coughing sputum, fever, chest pain and difficulty breathing). However, it is a fastidious bacterium that can be difficult to grow through culture. The clinical presentation of the patient makes the iSEP-SEQ result highly plausible. Without a clear microbiological diagnosis, a clinician may choose to administer broad-spectrum treatments, increasing the risk of AMR. This finding highlights the benefits of the iSEP-SEQ approach in accurately identifying the pathogen, enabling optimized treatment for life-threatening bloodstream infections, and potentially reducing risk of AMR.

While these results would benefit from confirmation by PCR, it highlights the potential of the microfluidic-enabled portable workflow for the culture-free and fast identification of sepsis-causing pathogens.

## DISCUSSION

While cfDNA has been shown to be a powerful biomarker, cfDNA extraction remains a challenge. Skilled staff are employed to process samples for cfDNA manually or using high-throughput robots. The former is subject to human performance and suffers from variability from one person to the next. The latter requires high-capital investment in centralised facilities, extensive staff training, high-maintenance and long turnaround time due to the need to batch processes. Thus, complex sample processing is a factor that restricts the clinical availability and time-to-result of cfDNA-based assays. This is especially relevant to a context like infection diagnosis where time is critical and where diagnosis may be needed in remote settings without high-capital infrastructure.

We have developed an integrated single-use cartridge produced by high-volume injection moulding and containing pre-filled reagents. Together with its companion automation platform, the device transforms a sample of plasma into an eluate of cfDNA in less than 45 minutes, with scope to further reduce the extraction time. In total, over 400 human plasma samples were used in the investigation of various parameters, including the elution buffer volume, mixing time and silica membrane lot number, as well as plasma and DPBS sample volume. A subset of these results has been presented here, and the complete dataset is made available. Following preliminary optimisation, the average recovery of CNASafe cartridge was found to be 100.5% relatively to the gold standard in the field of cfDNA extraction (QIAamp CNA kit, manual extraction), highlighting the performance of the proposed platform for rapid, automated, and highly efficient extraction of cfDNA from human plasma samples. It is difficult to compare to other integrated microfluidic platforms described in the literature as those did not compare their results to the same reference. CNASafe provides an automated solution for the extraction of cfDNA in an environment where speed is critical, and samples cannot be batched for delayed extraction.

While nanopore sequencing has been widely used for genomic surveillance of emerging viruses, its clinical metagenomic applications for pathogen detection remain limited, in part because of the complex sample preparation and the need for skilled staff to operate complex batched workflows. Here we demonstrated that the CNASafe cartridge reduces the time to identification of circulating microbial cfDNA without the need to batch samples. The sample preparation is pathogen-agnostic and does not introduce biases to organisms, unlike several methods that enrich pathogen-specific reads and focus on bacteria and fungi. Most metagenomic studies rely on Illumina sequencing platforms, which have sequencing run times exceeding 16 hours. In contrast, our sequencing run times did not exceed 6h on the low capital investment and portable ONT platform, and 2h sequencing may be sufficient to detect pathogens in circulation and responsible for diseases and infections (12,31). If reduced to 2h, the whole iSEP-SEQ workflow could be performed in under 5h, providing a field-based, fast, sequencing-based identification of blood-borne pathogens. We have demonstrated that the CNASafe cfDNA eluate is compatible with ONT sequencing, known to be sensitive to contaminants, and applied the complete workflow on ten clinical samples, uncovering several organisms which were not identified by blood cultures.

The proposed iSEP-SEQ workflow would enable prompt aetiological investigations in decentralised settings or clinics without well-equipped centralised laboratories, leading to more consistent, more accurate and faster results than current solutions, alleviating the need for additional invasive procedures (e.g., bronchoscopy or bronchoalveolar lavage), reducing the time spent in hospital and associated costs, as well as reducing adverse effects from broad-spectrum antibiotics, thus leading to better outcome for patients.

## Data Availability

All data produced in the present study are available upon reasonable request to the authors

https://figshare.com/s/3b1f2840e2000bd44eef

## AUTHORS CONTRIBUTIONS

The corresponding author takes full responsibility that all authors on this publication have met the following required criteria of eligibility for authorship: (a) significant contributions to the conception and design, acquisition of data, or analysis and interpretation of data; (b) drafting or revising the article for intellectual content; (c) final approval of the published article; and (d) agreement to be accountable for all aspects of the article thus ensuring that questions related to the accuracy or integrity of any part of the article are appropriately investigated and resolved. Nobody who qualifies for authorship has been omitted from the list.

**Conceptualization:** Linda Marriott, Ana Martinez Lopez, Antonio Liga, Nicholas Feasey, Shevin T. Jacob, Jamie Rylance, and Maïwenn Kersaudy-Kerhoas.

**Data curation:** Linda Marriott, Ana Martinez Lopez, Nicholas Feasey, Shevin T. Jacob, Jamie Rylance, and Maïwenn Kersaudy-Kerhoas.

**Formal analysis:** Linda Marriott, Ana Martinez Lopez, Radhe Shantha Kumar, Benjamin J. Parcell, Nicholas R. Leslie, Nicholas Feasey, Jamie Rylance, and Maïwenn Kersaudy-Kerhoas.

**Funding acquisition:** Nicholas Feasey, Shevin T. Jacob, Jamie Rylance, and Maïwenn Kersaudy-Kerhoas.

**Investigation:** Linda Marriott, Ana Martinez Lopez, Antonio Liga, Shevin T. Jacob, Jamie Rylance, and Maïwenn Kersaudy-Kerhoas.

**Methodology:** Linda Marriott, Ana Martinez Lopez, Antonio Liga, Kazuhiro Horiba, Amanda Warr, Shevin T. Jacob, Jamie Rylance, and Maïwenn Kersaudy-Kerhoas.

**Project administration:** Linda Marriott, Antonio Liga, Jamie Rylance, and Maïwenn Kersaudy-Kerhoas.

**Resources:** Jacob N. Phulusa, Laura Carey, Jamie Rylance, and Maïwenn Kersaudy-Kerhoas.

**Software:** Linda Marriott, Ana Martinez Lopez, Antonio Liga, Kazuhiro Horiba, and Yoshinori Ito.

**Supervision:** Yoshinori Ito, Jamie Rylance, and Maïwenn Kersaudy-Kerhoas.

**Validation:** Linda Marriott, Ana Martinez Lopez, Jamie Rylance, and Maïwenn Kersaudy-Kerhoas.

**Visualization:** Linda Marriott, Antonio Liga, and Maïwenn Kersaudy-Kerhoas.

**Writing - original draft:** Linda Marriott, Antonio Liga, and Maïwenn Kersaudy-Kerhoas.

**Writing - review & editing:** Linda Marriott, Ana Martinez Lopez, Antonio Liga, Kazuhiro Horiba, Amanda Warr, Jacob N. Phulusa, Radhe Shantha Kumar, Laura Carey, Yoshinori Ito, Benjamin J. Parcell, Nicholas R. Leslie, Nicholas Feasey, Shevin T. Jacob, Jamie Rylance, and Maïwenn Kersaudy-Kerhoas.

## CONFLICTS OF INTEREST

AL, MKK and KH have filed and been granted patents related to this work.

## ACKNOWLEDGEMENTS

We acknowledge Engineering and Physical Science Research Council EP/R00398X/1, Royal Academy of Engineering FoD scheme, Heriot-Watt University, Scottish Enterprise, Santander Universities Santander X award for funding.

## Notes

### Competing Interest Statement

Antonio Liga and Maiwenn Kersaudy-Kerhoas have granted and pending patents on elements of the cartridge design. Kazuhiro Horiba has a granted patent on the bioinformatic tool Pathdet.

### Funding Statement

This study was funded by the UK Research and Innovation Engineering and Physical Research Council EP/R00398X/1; the Royal Academy of Engineering FoDSF 1920 2 100018; Internal funds from Heriot-Watt University B19G1243; Scottish Enterprise Natantis HGSP Phases 1-2; Santander Universities - Santander X Award 2023

### Author Declarations

Research Ethics Committees Northern Ireland (ORECNI) of UK NHS gave ethical approval for part of this work (Reference: 18/NI/0148) National Health Sciences Research Committee of Malawi gave ethical approval for part of this work (Reference: 2157) Research Ethics Committee of Liverpool School of Tropical Medicine gave ethical approval for part of this work (Reference: 18-062) Research Ethics Committee of Heriot-Watt University gave ethical approval for part of this work (Reference: 1285 -iSEP-SEQ_ARCS / 320361)

