## Supplementary material for "An Automated and Portable Platform for Rapid Cell-Free DNA Isolation and Its Application in Microbial DNA Sequencing from Human Blood Samples"

**SUPPLEMENTARY MATERIALS**

**Supplementary Table 1**

| **Sample #** | Epi2me Data | | | | | | | | | | |
| --- | --- | --- | --- | --- | --- | --- | --- | --- | --- | --- | --- |
|  | Total Reads (all) | Reads failed Qscore filter (Q7) | Reads Analysed | Reads Classified | Reads Unclassified | % unclassified (unclassified/Analysed)*100 | Ave Seqence Length | Ave Quality Score | Cumulative Reads Homo sapiens | Identified organisms  (Species level, 3 reads or more) | |
|  |  |  |  |  |  |  |  |  |  | Name | No. of Reads |
| **1** | 957,362 | 64,471 | 892,890 | 880,089 | 12,801 | 1.4 | 506 | 11.41 | 880,050 | - | - |
| **2** | 230,872 | 23 | 212,297 | 209,362 | 2,935 | 1.4 | 468 | 11.44 | 209,324 | Human gammaherpesvirus 8 |  |
| **4** | 84,043 | 7,755 | 76,286 | 72,516 | 3,770 | 4.9 | 473 | 10.93 | 72,386 | Human gammaherpesvirus 8  Arcobacter cryaerophilus  Rhizobacter gummiphilus  Cutibacterium acnes | 6  4  3  3 |
| **5** | 47,082 | 2,682 | 44,398 | 43,088 | 1,310 | 3.0 | 519 | 10.85 | 43,073 | - | - |
| **6** | 209,971 | 14,073 | 195,898 | 176,554 | 19,344 | 9.9 | 401 | 10.66 | 175,068 | Methylobacterium sp. XJLW  Methylobacterium phyllosphaerae  Methylobacterium oryzae | 178  136  118 |
| **7** | 292,367 | 18,937 | 273,425 | 265,775 | 7,650 | 2.8 | 514 | 11.42 | 265,679 | Cutibacterium acnes  Flavobacterium sp. KBS0721  Methylobacterium sp. XJLW  Proteus mirabilis | 5  3  3  3 |
| **8** | 321,387 | 32,131 | 289,255 | 283,976 | 5,279 | 1.8 | 550 | 10.61 | 283,311 | Streptococcus pneumoniae  Methylobacterium phyllosphaerae  Methylobacterium sp. XJLW | 517  9  9 |
| **9** | 222,499 | 19,327 | 203,172 | 200,354 | 2,818 | 1.4 | 525 | 11.2 | 200,332 | Methylobacterium sp. XJLW | 3 |
| **10** | 386,144 | 20,540 | 365,604 | 360,640 | 4,964 | 1.4 | 459 | 12.46 | 360,512 | Methylobacterium phyllosphaerae  Methylobacterium sp. XJLW  Methylobacterium oryzae | 26  22  9 |

Supplementary Table 2: Statistics for EPI2ME bioinformatic workflow

**Supplemental Method 1**

The fluidic channels (which are featured in both sides of the fluidic side) were closed by welding of a thin COC layer to each side of the cartridge. Furthermore, the fluidic side is fitted with needle-like structures (1.5 mm OD, 0.8 mm ID, 1.8 mm height) which allow the piercing of the front face foil of the reagent side, thus opening the fluidic path within the cartridge. Placed around each of these piercing structures is a soft ring which guarantees fluidic sealing while the two sides are pressed together. These O-rings are toroids derived by punching a 1mm 30˚ silicon layer (Silex ltd) with a 3mm and a 1.5 mm biopsy punch.

The reagent side of the CNASafe cartridge was prepared by drilling two 1.6mm filling holes into each reagent chamber before the front face was sealed with LINC 1298 Heat Seal Foil (Label Innovation). Sealing was carried out by a third party until 12/05/2021, after which sealing was carried out in-house. Reagent chambers were filled with proprietary extraction reagents and sealed with QuickSeal Foil PCR Ultra™ Self Adhesive Sealing Film (IST Scientific) prior to experiments. The optimisation of elution buffer volume reported in the Results section was performed prior to the sealing being carried out in-house. All other results were carried out after sealing was done in-house.

The custom alignment frame feature was designed with the multiple aims to 1) aligning the two sides of the cartridge 2) avoiding unwanted piercing during transportation and 3) keeping the parts together after the CNA extraction to avoid potential spillage of liquid in the case of incorrect emptying of the cartridge. The two cartridge sides are initially separated by the alignment feature, which is meant to allow safe transport of the cartridge maintaining the two parts connected but separated.

The membrane (Mini Spin column (Qiagen)) was prepared by piercing a small hole in the lid of the column and positioning rigid tubing (ID 0.8 mm, length 75 mm) just above the membrane. The tubing was secured into place and the lid of the column sealed using epoxy glue (Loctite 3430, Henkel). All prepared columns were pressure-tested prior to use for quality control. Only columns with a determined dry baseline pressure drop of 7.4 +/- 1.1 kPa were used. The external membrane along with tubing for the connection to the sample tube, was attached to the fluidic side before the two sides of the cartridge (reagent and fluidic) were loaded onto a custom automation platform. Here the two sides are brought together with the aid of an alignment frame, resulting in the piercing of the front face foil (of the reagent cartridge) by the needle-like projections on the fluidic side.

**Supplemental Method 2**

qPCR analysis was performed using StepOne™ Software v2.1. Outliers were identified as having a Cq valve of >0.3 between technical replicates. Cq values of NTC were above 30 with average Cq values for standards ranging from 10.17 (10ng/µL) to 20.92 (0.01ng/ul), resulting in an average r^2^  of 0.997 across runs.

**Supplemental Figure 1**

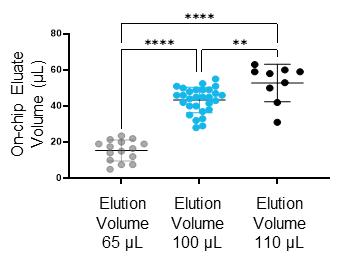

*Supplementary Figure 1: Impact of on-chip elution volume on eluate volume.*

**Supplemental Figure 2**

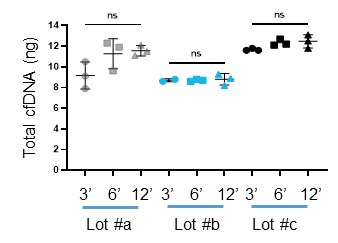

*Supplementary Figure 2: Impact of lot number and mixing time on absolute amount of cfDNA (ng).*

**Supplemental Figure 3**

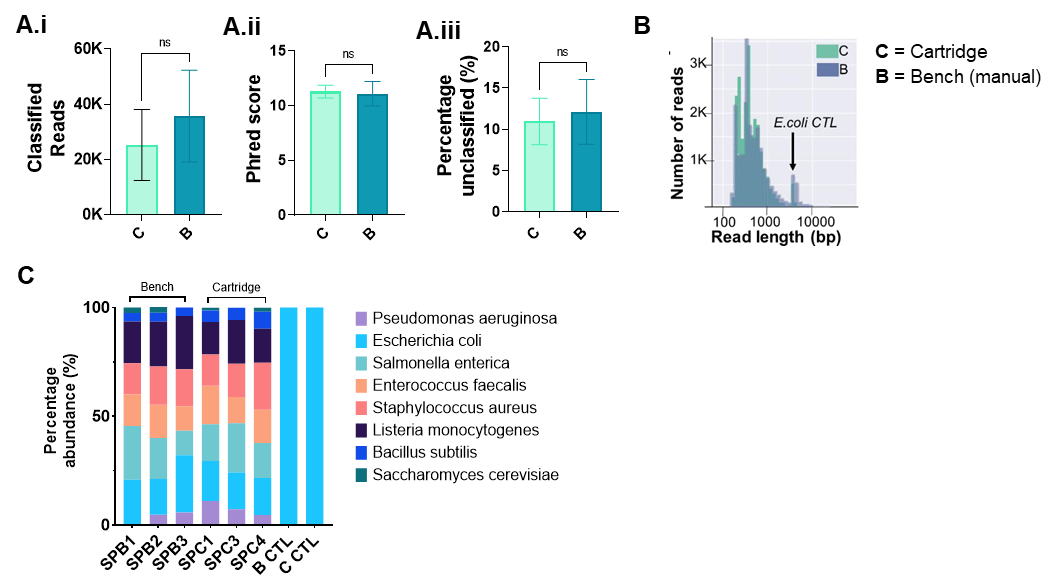

*Figure S3: Characterisation of sequencing workflow on Cartridge (C and Benchmark eluates.* ***(A)*** *Comparison of Classified reads (A.i), Phred score (A.ii) and Percentage of unclassified reads (A.iii) between cartridge (C) or bench (B) extractions (N=3).* ***(B)*** *Comparison of number of reads vs read lengths for Cartridge and Bench eluates* ***(C )*** *Relative abundance of microbial mix in bench (SPB1-3) and cartridge (SPC1, 3-4) extracted eluates for each of the 8 microbe genomic DNA included in the community mix. Plasma only controls on Bench and Cartridge (B CTL and C CTL).*
